# Characterizing Dementia Phenotypes from Unstructured EHR Notes with Generative AI and Interpretable Machine Learning

**DOI:** 10.1101/2025.10.01.25336815

**Authors:** Alice S. Tang, Billy Z.D. Zeng, Katherine P. Rankin, Bruce Miller, Maria Luisa Gorno-Tempini, William W. Seeley, Howard J. Rosen, Gil D. Rabinovici, Tomiko T. Oskotsky, Marina Sirota, Pedro Pinheiro-Chagas

## Abstract

Dementia encompasses diverse clinical syndromes where diseases of the brain can manifest as impaired cognitive abilities, such as in Alzheimer’s disease (AD) and behavioral-variant frontotemporal dementia (bvFTD). The diversity of symptom presentations often results in challenges in diagnosis. Crucial clinical information remains in unstructured narrative notes within electronic health records (EHRs). We leverage large language models (LLMs) for symptom phenotyping from notes in UCSF Information Commons, focusing on patients with expert dementia syndrome diagnosed from a multidisciplinary team of specialists from the UCSF Memory and Aging Center. We developed a pipeline to extract findings in a validated structured output, clustered into symptom groups, and then classified patients into syndromes with traditional machine learning paradigms. From over 9,000 cross-referenced patients and over 350,000 specialty-related notes, matched cohorts of bvFTD (122 patients) and AD (170) syndromes were identified. From notes, 12,637 distinct symptom phrases were extracted, with clustering analysis revealing 51 symptom groups. A logistic regression model separated AD and bvFTD with an AUC of 0.83. Disinhibition and obsessive-compulsive behaviors favored bvFTD, while anxiety and visuospatial abnormalities favored AD. This novel approach, combining LLM-based structured information extraction with traditional interpretable prediction paradigms, demonstrates a promising approach for enhanced symptom characterization in dementia. Our findings suggest potential future applications in improving diagnostic accuracy, developing prediction models, and optimizing treatment strategies in dementia care.

## Introduction

Dementia refers to a neurodegenerative process that leads to deficits in cognitive function, which may encompass heterogeneous clinical syndromes including Alzheimer’s disease, vascular dementia, Parkinson’s disease, and frontotemporal dementia^1,2^. The varied and heterogeneous presentation includes symptoms that overlap across many medical domains, with neurological and psychiatric manifestations that ultimately may lead to a work-up that results in a neurodegenerative diagnosis. Yet more than 50% of patients do not receive specialized neurologic care for a diagnosis in primary care, which can lead to delays and misdiagnosis until moderate or advanced stages^3–5^. Furthermore, the symptoms and clinical manifestations do not predict neuropathological diagnosis, such as abnormal protein deposition that leads to Alzheimer’s disease and frontotemporal dementia^6–8^. The biological processes are ultimately important to identify targets and eligible patients for drug discovery and early intervention. As such, there is a need for improved early identification of clinical syndromes and connection with mechanisms in order to intervene early and accurately before irreversible decline.

Recent advancements in large clinical datasets have enabled exciting opportunities and advances around understanding neurodegeneration, especially with the availability of rich longitudinal data. Electronic health records (EHRs) in particular have become a valuable source of real-world clinical data, offering structured information that provides useful insights into the heterogeneity of dementia over time. Multiple prior works leveraging structured data to diagnose and predict AD and neurodegeneration heterogeneity^9–11^, though limitations exist^12^. Most notably, much of the essential clinical detail remains embedded within unstructured narrative notes, including detailed free-text documentation that captures detailed clinical observations, nuanced symptom descriptions, and disease trajectories often not captured by structured data, which often only includes billing or diagnostic codes and may be fraught with inaccuracies.

These vital details, reflecting the dynamic nature of dementia morbidities and corresponding support needs, are challenging to interpret from the vast amount of text. Furthermore, dementia treatment and management may involve multifactorial approaches, including medications, therapies, and social interventions specific to each patient^2,13^. Unlocking information in clinical notes is critical, as the significant overlap in neurological, psychiatric, and behavioral symptoms across different dementia syndromes, such as Alzheimer’s disease and frontotemporal dementia, may often manifest as subtle signs before overt cognitive deficits and neurological work-up^14,15^.

Traditional artificial intelligence, particularly interpretable machine learning models such as linear regression and decision tree-based methods, has been instrumental in advancing dementia research using structured data. These paradigms have been applied to various data modalities, including biomarkers^16,17^, genetics^18,19^, neuroimaging^20,21^, speech^22^, and large clinical datasets^23–25^. The core advantage of these models lies in their interpretability based on coefficients or weights in the models. For example, a model trained on structured clinical data was able to predict Alzheimer’s disease onset risk while highlighting the predictive features, such as diagnoses that could inform discovery^10^. However, the performance of these models are constrained by the inherent limitations of structured data. Key information regarding a patient’s symptoms and cognitive state is often missing from structured fields and exists only in clinician notes, creating a gap in data that hinders both feature extraction and the power of predictive modeling. Large language models (LLMs) have also demonstrated remarkable capabilities in natural language understanding and concept extraction, offering a transformative potential to analyze these complex, unstructured clinical notes to improve prediction tasks. This has been particularly the case in applications within social determinants of health extractions from notes^26,27^. Within dementia, natural language processing has been utilized in prior work to extract specific domains of symptomatology of dementia from clinical notes^28,29^. Our previous work employed machine learning on EHR and omics data to extract valuable clinical features for dementia subgroups^10^. Recent work includes exploration of agentic LLM workflow to extract cognitive concerns in dementia^30^, and LLM combined with traditional machine learning ensemble models to enhance early prediction of dementia^31^. These approaches have not been applied across a patient’s clinical care trajectory in dementia.

In this work, we leverage generative AI (LLMs) on a large number of patients, to analyze unstructured clinical data with well-phenotyped neurodegenerative diagnoses from expert neurologists from the University of California San Francisco (UCSF) Memory and Aging Center. We employ a novel agentic workflow, using specialized LLMs for a sequence of tasks, including data preprocessing, prompt tuning, and evaluation. In particular, our main novelty includes the combination of generative AI and traditional ML approaches. We demonstrate a proof of concept with well-phenotyped Alzheimer’s disease and behavioral variant frontotemporal dementia (bvFTD), demonstrating how symptom extraction can aid with discovery science and disease classification.

Our work builds upon previous work by both helping to derive specific clinical features and symptoms of dementia and using an LLM approach for prediction. Ultimately, this structured data could form the basis for a clinical decision support tool designed to help physicians improve diagnostic reasoning, referrals, or ultimately improve care through summaries and symptom highlighting, potentially a multidisciplinary approach to neurodegenerative diseases and refining patient care in this field.

## Methods

### Cohort Identification and Filtering

The UCSF Information Commons captures EHR data from over 5 million patients. Patients were cross-referenced based on presence in the UCSF Memory and Aging Center research database to select patients with expert diagnoses. Notes from these patients were then extracted.

Given the heterogeneity of notes, a hierarchical filtering strategy was applied, focusing on neurodegeneration-relevant specialties. Specifically, notes from neurology-related specialties (e.g., Neurology, Neuropsychology, Neurosurgery, etc.) and support providers (e.g., Speech and Language Pathologists) were identified from all available visits, irrespective of time. To optimize cost efficiency and account for variations in note context, a local LLM was utilized to pre-filter notes for the presence of extractable information. Extractable information was defined in a custom prompt to assess each clinical note for diagnostic codes (e.g., ICD, DSM), clinical findings (symptoms, physical exam), measurements (cognitive tests), medications, or medical history. We utilized a local LLAMA3.3 model with a controlled (zero temperature) setting to generate deterministic outputs.

Furthermore, a structured output schema was enforced using a Pydantic model, ensuring outputs follow a JSON format with boolean indicators for each clinical element. The AI-generated responses were then validated against this predefined schema for consistency. From this extraction process, a refined dataset of notes containing at least one extractable piece of an information was filtered for subsequent detailed extraction.

### Selecting Patients with AD and bvFTD

Alzheimer’s disease and behavioral variant frontotemporal dementia were chosen for this study, given the clinical syndromes can present with overlapping presenting cognitive and behavioral features, particularly for younger patients^32^. Moreover, there are now FDA-approved disease-modifying treatments for Alzheimer’s disease, and distinguishing from other neurodegenerative diseases can better inform patient selection for improved specificity when evaluating for treatment effects in clinical trials or in care. In order to adjust for hospital utilization and note characteristics, we utilized propensity score matching between AD and bvFTD patients with a nearest-neighbor matching strategy. Matching was performed on note metadata (minimum note length, log(average note length), log(number of notes), minimum year, duration of notes) and presence of clinical element (diagnostic code, clinical finding, measurements, medication, medical history) based on LLAMA output, with natural log representation for log.

### Leveraging LLM for Information Concept Extraction

To extract detailed unstructured information from notes, we employed a structured prompt with GPT-4o from Microsoft Azure OpenAI service (gpt-4o-2024-08-06 model) with a temperature setting of 0 on the filtered and selected clinical notes. The prompt is available in **Supplement**. This is done through the UCSF Versa API, which is available as part of the UCSF Versa Generative AI Platform, an enterprise platform designed to enable clinicians, healthcare operators, and researchers access to secure generative AI tools. UCSF Versa is approved for use with protected clinical data and has undergone security assessments to ensure that data only remains within the UCSF environment. The API allows a secure gateway to generative AI deployments and enables the incorporation of generative AI in data pipelines.

For information extraction, a predefined structured JSON output schema was enforced with Pydantic. The extracted detailed clinical information is structured into the following domains:

- Diagnostic Codes: Complete diagnostic codes, source manuals (ICD-9, ICD-10, DSM-IV, DSM-5), diagnosis terms, status, supporting evidence, and confidence levels.
- Clinical Findings: Specific clinical symptoms/signs, predefined neurological/ neuropsychiatric categories, temporal details, severity, supporting evidence, and confidence levels.
- Structured Measurements: Measurement values, units, test types, test names, cognitive/behavioral domains, supporting evidence, and confidence levels.
- Medications: Medication names, dosages, frequencies, statuses, supporting evidence, and confidence levels.
- Medical History: Documented conditions, onset details, status, treatment history, supporting evidence, and confidence levels.

We prioritized clinical findings given the unlikelihood of symptoms and signs being present in structured data. This is attributable to the absence of an ontology, billing codes, or other standardized methods for structured recording within clinical workflows. Consequently, clinical findings hold the greatest significance for the extraction and augmentation of structured data.

### Symptom Text Processing and Embedding

Clinical findings were extracted based on the ‘supporting evidence’ field, which contains verbatim text excerpts from clinical notes. The extracted text was then preprocessed using a custom NLP function to: convert lists or non-string types to a uniform string format, normalize text by converting Unicode characters to ASCII, lowercase the text for consistency, remove non-alphanumeric characters (excluding slashes, hyphens, and whitespace), and eliminate isolated single consonants and extra whitespace.

Each cleaned clinical finding text was then individually embedded using the Microsoft Azure OpenAI platform (text-embedding-3-large-1 model) through UCSF Versa API, with resulting vector representations in 3072 dimensions. The embeddings were further reduced to 100 dimensions using principal component analysis (PCA) for computational efficiency and noise reduction. For visualization purposes, UMAP was applied to the PCA embeddings to project into 2 dimensions.

### Symptom Clustering and Labelling

For clustering of embedding extracted clinical findings, we utilized HDBscan on the UMAP-projected data with grid-based parameter optimization for min_cluster_size (50, 75, 100, 200), min_samples (5, 10, 15, 20), and cluster_selection_epsilon (0, 0.05, 0.1), to determine the optimal clustering configuration. With HDBscan, some data can be labeled as noise and removed from downstream analyses.

For each identified cluster (with noise excluded), 15 text examples were randomly selected from the “supporting evidence” field, which contains the verbatim clinical text for the relevant symptom cluster. These examples were then provided to GPT-4o (model: gpt-4o-2024-08-06, temperature set to 0) via UCSF Versa API to automatically generate concise, descriptive labels for each cluster. These labels were then parsed and aggregated into a structured JSON output containing cluster IDs and labels. Top symptom clusters were then investigated between neurodegenerative syndromes.

### Patient Representation and Classification

With 51 symptom clusters to help represent patient clinical findings, each patient was represented based on the number of counts of findings corresponding to each cluster. Generated labels were integrated for clarity. Patients were then split into 80% training and model optimization set, and 20% final evaluation set. A regularized logistic regression with a stratified K-fold cross-validation framework was utilized on patient representations to assess the classification of AD or bvFTD.

Model evaluation includes receiver operating characteristic (ROC) area under the curve (AUC), confusion matrices, and precision-recall curves based on evaluation set performance. To optimize interpretability for the model and enable insights, we applied recursive feature elimination with cross-validation (RFECV) and permutation testing to assess stability and significance, and we retrained the model with the optimal number of cluster groups with re-evaluation on the evaluation set. Model coefficients and permutation importance scores were investigated to identify key symptom clusters associated with the prediction of each neurodegenerative syndrome for key insights. Permutation importance was computed on the whole dataset based on the impact on AUC.

### Ethical Approval

The Institutional Review Board of University of California San Francisco gave ethical approval for this work (IRB #20-32422).

## Results

### Local language model filters notes with extractable concepts

From the UCSF EHR database, 9169 patients cross-referenced with the UCSF Memory and Aging Center research database were identified. After filtering patients to those with neurology, psychology, and psychiatry-related notes, there were 4,728 patients with a combined 358,816 clinical notes (details in **Methods, Supplement**, and **Figure 1**). To optimize energy use and cost of LLM models^33^, we screened notes with a locally hosted LLM model to determine the availability of extractable information, especially clinical findings. After filtering based on extractable information determined by the local LLM, 4,690 and 61,823 notes remained (**Figure 2A**).

**Figure 1:**
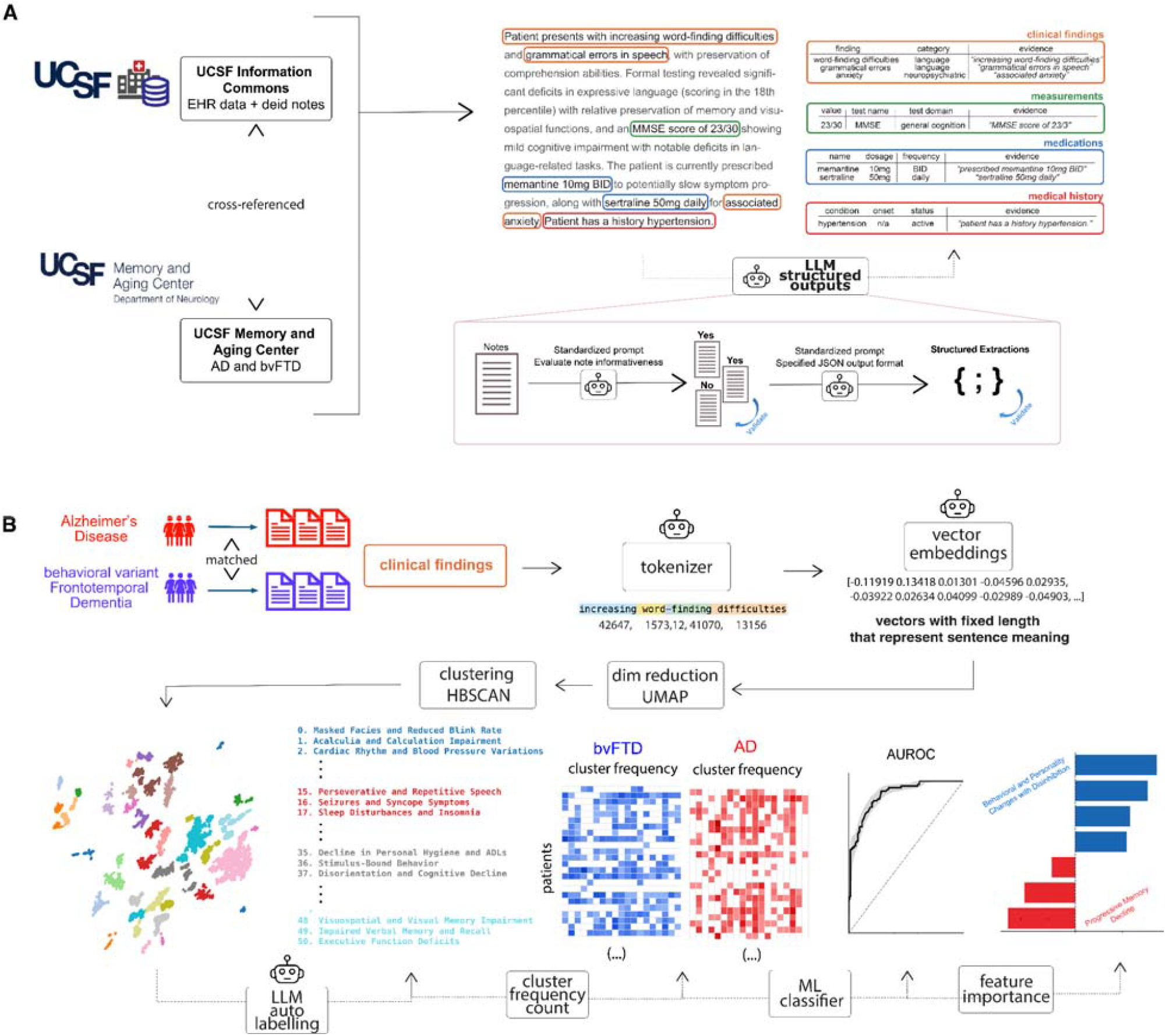
Overview of Dementia Structured Output Extraction and Application. A. Data source from UCSF Information Commons cross-referenced with expert diagnosis from the UCSF Memory and Aging Center. Example extraction of LLM structured outputs. B. AD and bvFTD were chosen for proof of concept for symptom phenotyping. Patients were matched based on note characteristics, and clinical findings were extracted. These findings were subsequently embedded and clustered to identify symptom cluster groupings, which are also labelled with an LLM. Patients are then labelled based on symptom group membership, and models were trained for classification and interpretation.

**Figure 2.**
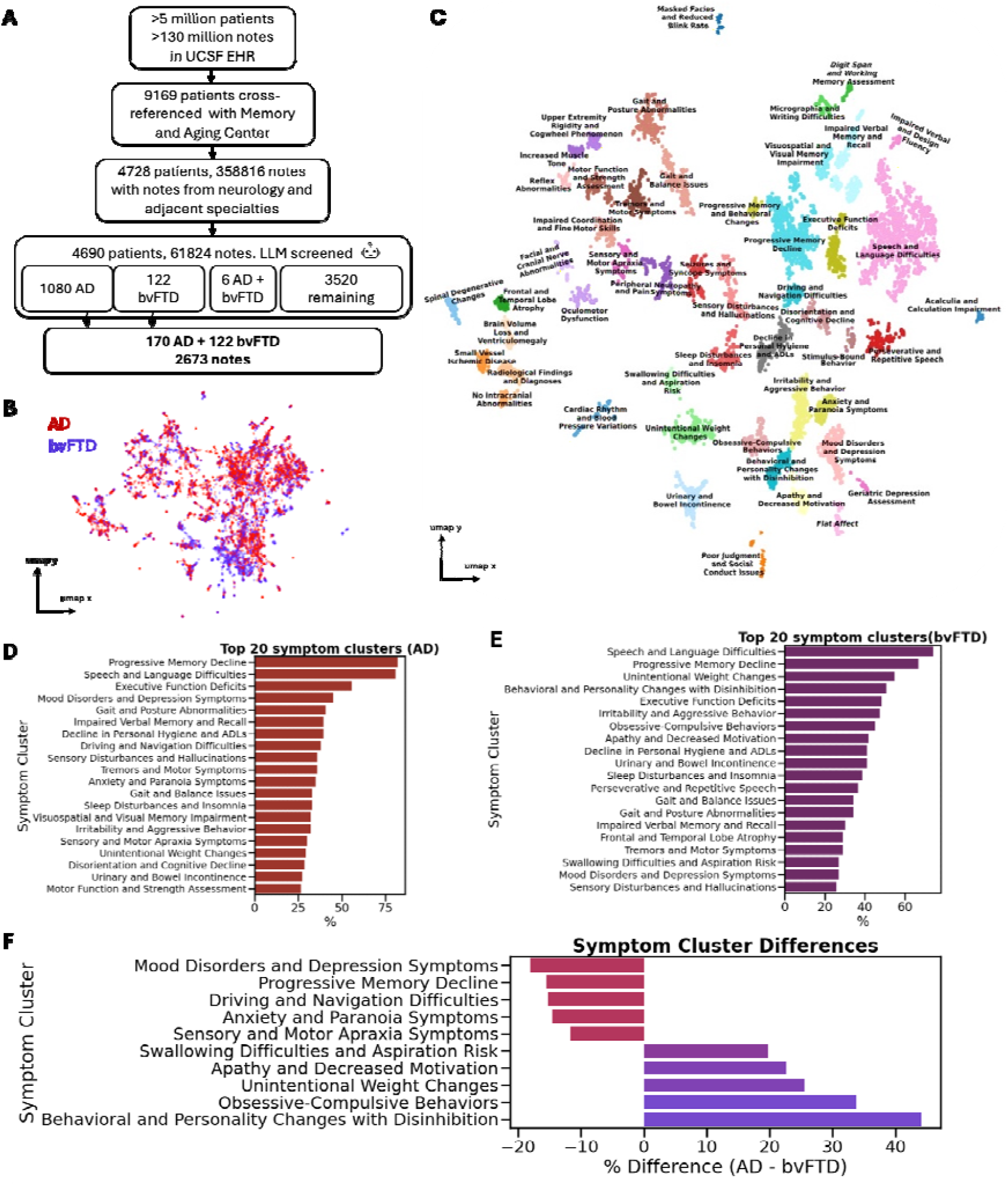
Structured LLMs extractions enable symptom phenotyping in AD and bvFTD. A. Attrition and patient flow from the large EHR database to the patients evaluated, with matched AD and bvFTD below. B. 2D umap representation of extracted clinical findings, embedded with OpenAI text embedding model. C. Symptom clusters identified with HDBScan, labelled with LLM D. Top symptom clusters in the Alzheimer’s disease group E. Top symptom clusters in the behavioral variant Frontotemporal Dementia group F. Symptom cluster proportion differences between AD and bvFTD

From these, 122 bvFTD patients and 1,080 AD patients were identified. To account for potential confounding, we identified propensity-score matched cohorts of 122 bvFTD and 170 AD patients balanced on per-patient filtered note characteristics, resulting in a total of 292 patients and 2,673 notes. After matching, note characteristics are balanced (**Table 1**).

**Table 1:**
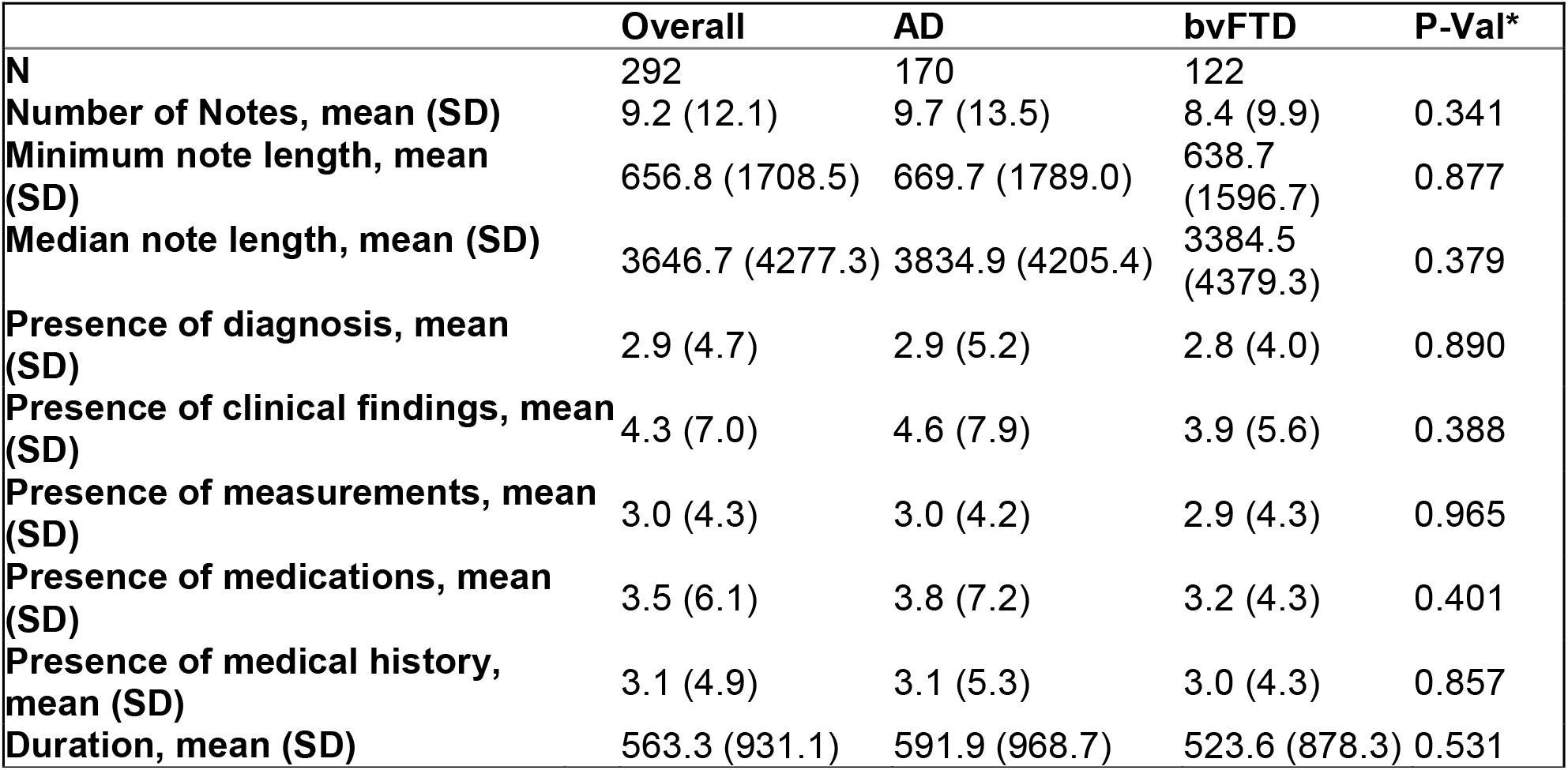
AD and bvFTD patient note characteristics. N represents number of patients, while duration represents the days between the patient’s first and last note. Note characteristics are otherwise aggregated for each patient.

|  | **Overall** | **AD** | **bvFTD** | **P-Val\*** |
| --- | --- | --- | --- | --- |
| **N** | 292 | 170 | 122 |  |
| **Number of Notes, mean (SD)** | 9.2 (12.1) | 9.7 (13.5) | 8.4 (9.9) | 0.341 |
| **Minimum note length, mean (SD)** | 656.8 (1708.5) | 669.7 (1789.0) | 638.7 (1596.7) | 0.877 |
| **Median note length, mean (SD)** | 3646.7 (4277.3) | 3834.9 (4205.4) | 3384.5 (4379.3) | 0.379 |
| **Presence of diagnosis, mean (SD)** | 2.9 (4.7) | 2.9 (5.2) | 2.8 (4.0) | 0.890 |
| **Presence of clinical findings, mean (SD)** | 4.3 (7.0) | 4.6 (7.9) | 3.9 (5.6) | 0.388 |
| **Presence of measurements, mean (SD)** | 3.0 (4.3) | 3.0 (4.2) | 2.9 (4.3) | 0.965 |
| **Presence of medications, mean (SD)** | 3.5 (6.1) | 3.8 (7.2) | 3.2 (4.3) | 0.401 |
| **Presence of medical history, mean (SD)** | 3.1 (4.9) | 3.1 (5.3) | 3.0 (4.3) | 0.857 |
| **Duration, mean (SD)** | 563.3 (931.1) | 591.9 (968.7) | 523.6 (878.3) | 0.531 |

### LLM extracts groups of findings from unstructured clinical notes

With UCSF Versa API (GPT4o), we extracted 12,637 distinct clinical findings, with an average of 43 findings per patient. UMAP of the embeddings of verbatim symptom text excerpts are shown in **Figure 2B**, and 51 symptom clusters were identified with an optimal silhouette score of 0.549 (**Figure 2C**). Approximately 15–20% of data points were labeled as noise. As an example, the cluster “Acalculia and Calculation Impairment” includes phrases such as ‘*difficulty solving mathematical calculations’*, ‘*missing points on a complex multiplication problem’*, and ‘*trouble with calculations, which is highly unusual’*. **Supplemental Table 1** shows 6 example symptom clusters with example text.

When directly comparing concept groups between the the two syndromes, patterns in symptom cluster groups emerge. The top 5 AD symptom clusters include progressive memory decline, speech and language difficulties, executive function deficits, mood and depression symptoms, and gait and posture abnormalities. The top 5 bvFTD symptom clusters include speech and language difficulties, progressive memory decline, unintentional weight changes, behavioral and personality changes with disinhibition, and executive function deficits (**Figure 2D, 2E**). Distinct and shared symptom clusters also exist. Shared symptoms include progressive memory decline and speech and language difficulties. Mood and depression symptoms stand as most prominent among AD, while behavioral and personality changes with disinhibition stand as most prominent among bvFTD (**Figure 2F, Supplement**).

### Traditional ML can classify dementia syndrome

Patients represented by number of mentions for each symptom cluster, which were then added as inputs to the classification model. For the goal of interpretation, we performed RFECV, identifying an optimal number of 39 features that maximized cross-validation AUC (**Figure 3A**). Symptom clusters that were selected with 100% frequency include: anxiety and paranoia symptoms, apathy and decreased motivation, disorientation and cognitive decline, behavioral and personality changes with disinhibition, geriatric depression assessment, stimulus-bound behavior, obsessive-compulsive behavior, oculomotor dysfunction, poor judgment and social conduct issues, micrographia and writing difficulties, and sensory and motor apraxia symptoms (**Figure 3B**). Symptom clusters that were utilized with 0% frequency include increased muscle tone, speech and language difficulties, sleep disturbance and insomnia, radiological findings and diagnoses, and upper extremity rigidity and cogwheel phenomenon.

**Figure 3.**
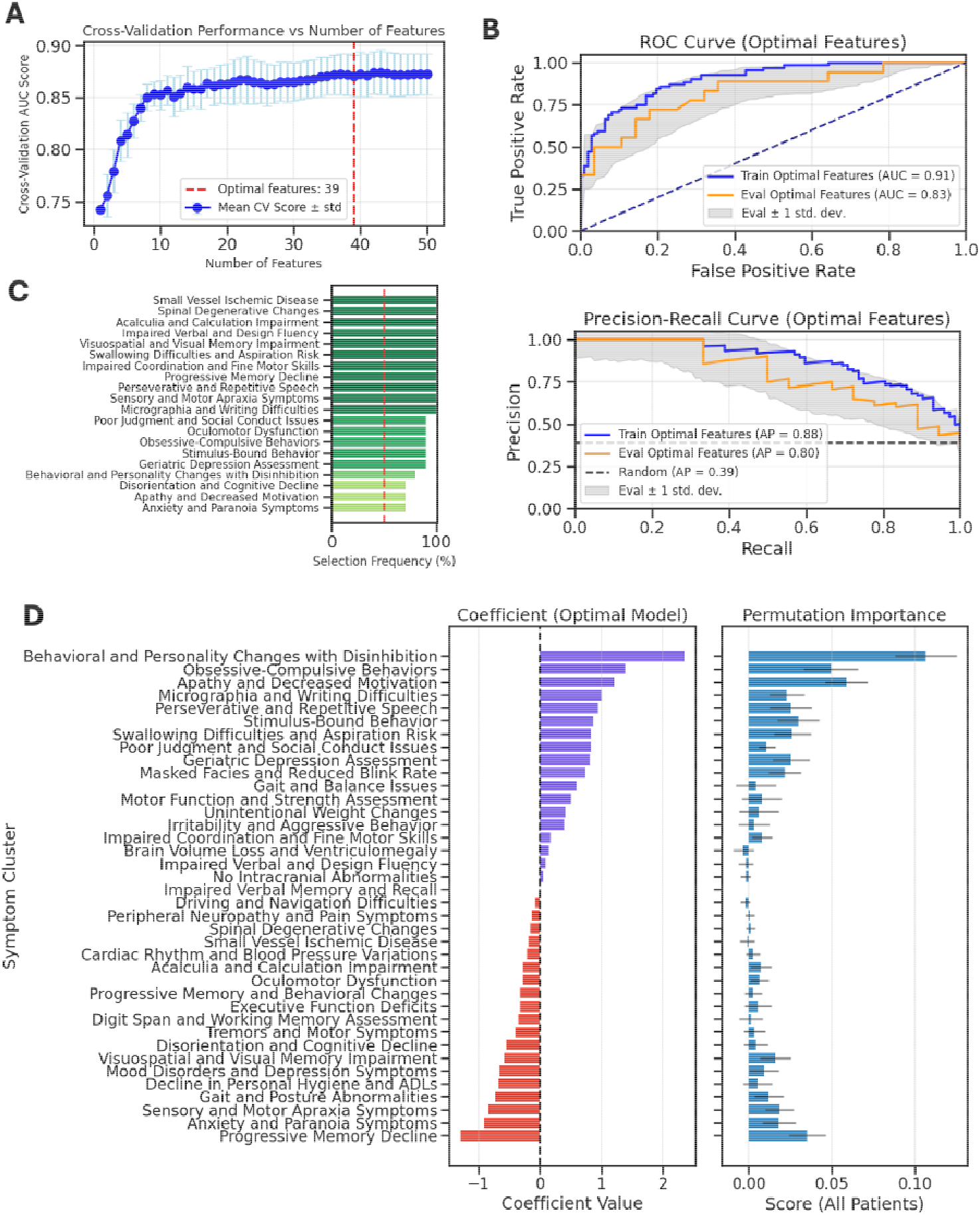
Traditional ML Classifies AD vs. bvFTD from Symptom Groups. A. Plot of the training sample AUC score based on the number of features in iterations of recursive feature elimination with standard deviations shown. The average optimal number of features is 39, represented by the red dotted line. B. Top 20 symptom clusters, based on utilization frequency during iterations of recursive feature elimination. Red line represents 50% utilization frequency. C. Performance of linear regression model with optimal features. ROC curve (top) and precision recall curve (botton) for the training sample (blue) and held-out evaluation sample (orange) with standard deviations from bootstrapped samples of the evaluation set (grey). D. Coefficients of each symptom cluster for the optimal feature logistic regression model (left), with positive coefficients favoring bvFTD (purple) and negative coefficients favoring AD (red). Robustness of feature importance was evaluated based on performance change with permutations of each symptom cluster, with AUC score changes and standard deviations shown.

With the optimal set of features, the retrained logistic regression model was performed on the evaluation set with AUC 0.833 ± 0.072 and average precision 0.793 ± 0.132 (**Figure 3C**). Model coefficients highlight symptom cluster importance for classifying bvFTD or AD syndromes. Symptom clusters that influence the model to predict bvFTD include behavioral and personality changes with disinhibition (with the highest coefficient value), obsessive-compulsive behaviors, apathy and decreased motivation, micrographia and writing difficulties, and perseverative and repetitive speech. Symptom clusters that influence the model to predict AD include progressive memory decline, anxiety and paranoia symptoms, sensory and motor apraxia symptoms, gait and posture abnormalities, decline in personal hygiene and ADLs, mood disorders and depression symptoms, and visuospatial and visual memory impairment (**Figure 3D**). Symptom clusters that do not contribute much to prediction include impaired verbal memory and recall, no intracranial abnormalities, driving and navigation difficulties, and impaired verbal and design fluency. Symptom cluster importance was also determined via permutation to determine the robustness of top features, supporting overall trends from model coefficients, with decreased importance assigned to some symptom clusters. Specifically, the influence of decline in personal hygiene and ADLs for AD prediction is no longer robust with permutation, while acalculia and calculation impairment and oculomotor dysfunction have relatively increased importance for AD. For bvFTD, irritability and aggressive behavior have relatively increased importance with permutation (**Figure 3E**).

## Discussion

Our study demonstrates the utility of LLMs in extracting and structuring symptom outputs from clinical notes to differentiate between dementia subtypes, through understanding of symptom differences given expert neurodegenerative diagnosis. This novel approach, combining LLM-based structured information extraction with traditional machine learning techniques, demonstrates promising results for enhanced symptom characterization and phenotyping in dementia. Our findings suggest potential applications in improving diagnostic accuracy, developing classification models, and optimizing treatment strategies in dementia care.

The identified symptom clusters extracted from clinical notes span a wide range of domains, including cognitive, behavioral, motor, autonomic functions, and imaging findings, many of which are not systematically captured by billing codes or standardized assessments. Symptom prevalence in AD and bvFTD revealed expected differences, such as a higher prevalence of memory decline in AD and more prominent behavioral and personality changes in bvFTD. However, these symptoms are common in both conditions and may complicate diagnosis during early presentations or represent the progression of disease^32,34^. This underscores the diagnostic challenge clinicians face and validates the need for data-driven methods that can weigh the importance of a comprehensive symptom profile. To address this challenge, our classification model identified the most informative symptom clusters for distinguishing between the two syndromes.

For bvFTD, the model assigned the highest importance to behavioral features, including disinhibition, obsessive-compulsive behaviors, apathy, and stimulus-bound behavior, which aligns with the core diagnostic criteria. Interestingly, the model also highlighted micrographia and writing difficulties, which aligns with findings that Parkinsonism features tend to be more prevalent in FTD (almost 40% prevalence)^35,36^ than AD (9.7% prevalence in AD without Lewy bodies)^37^. These findings suggest that a constellation of behavioral and specific motor symptoms is highly predictive of bvFTD relative to AD.

Conversely, the model indicated that an AD diagnosis is associated with more variable symptomatology. While progressive memory decline was an important predictor, the model also emphasized the significance of anxiety, paranoia, and depressive symptoms, supporting existing literature on the early and prominent psychiatric manifestations of AD^38–40^. Furthermore, our model highlighted the importance of visuospatial and sensorimotor deficits, including sensory and motor apraxia, gait abnormalities, and oculomotor dysfunction. This constellation of findings suggests that deficits in higher-order sensory processing and motor planning, beyond just memory, are key distinguishing features of AD in this context.

Notably, imaging features such as generalized brain volume loss or non-specific ischemic changes on imaging were not found to be highly discriminative in our model. This suggests that while these features are common in aging and neurodegeneration, they may not be specific enough to differentiate between AD and bvFTD pathology in the absence of more granular symptom data. This alludes to the importance of clinical exams as well as patient and collateral history in the differentiation and management of dementia^2^.

A key strength of our study is the use of highly-validated, expert-adjudicated diagnoses from the UCSF Memory and Aging Center, which serves as a gold standard for labeling clinical syndromes. This ground truth, combined with the analysis of longitudinal data from patient notes over time, allows for a more reliable and nuanced derivation of disease-specific symptoms compared to studies relying on billing codes or cross-sectional data alone.

Our study also has several limitations. The model was trained on data from a single academic medical center, and its performance may vary in other healthcare settings. Furthermore, as clinical documentation shifts from clinicians’ typed notes towards notes from AI scribes, salient features that are identified by classification models may change. AI scribe documentation may leave out important nuances that are present in clinician-typed notes^41,42^. LLM-based classifiers trained on clinician-authored notes may rely heavily on structured language, standardized phrasing, and implicit clinical reasoning that may not be present in documentation produced by AI. Consequently, the classification performance of LLM-based models may vary. Our classification models are also based on all neurology-related notes for a patient across the patient’s clinical trajectory. Future work will investigate notes from a wider range of specialties and the temporal relationships between extracted symptoms and dementia onset, as well as prediction across the temporal spectrum. Our work also focuses only on clinical symptoms, future work will incorporate structured data such as the patient’s medical history and medications that exist both in the EHR and extracted clinical notes. Nevertheless, our study demonstrates the utility of structured symptom extraction combined with expertly diagnosed dementia syndromes for phenotyping and classification.

## Conclusion

Here, we utilize an LLM-based structured information extraction approach leveraging GPT-4o and LLama 3 for enhanced symptom characterization and phenotyping in expertly diagnosed neurodegenerative diseases from the UCSF Memory and Aging Center. We present a novel agentic approach using specialized LLMs for a series of tasks, including data preprocessing, prompt tuning, and output evaluation. Furthermore, we illustrate, via a proof-of-concept on AD and bvFTD, the utility of symptom extraction in discovery science and disease classification. We envision this work as a foundational step toward a future clinical decision support tool that can assist clinical diagnostic reasoning and management, including decisions around specialized referrals and resources based on a patient’s specific symptom burden. Future endeavors will focus on characterizing symptoms across various dementia presentations and developing personalized approaches to dementia care.

## Data Availability

Access to EHR databases and participant-identifiable information are controlled due to the sensitive nature of the data. The UCSF EHR database can be accessed by UCSF-affiliated individuals by contacting UCSF Clinical and Translational Science Institute or UCSF Information Commons team. If the reader is unaffiliated with UCSF, they can set up an official collaboration with a UCSF-affiliated investigator by contacting the principal investigator, M.S. Participant data from the UCSF Memory and Aging Center can be requested at https://memory.ucsf.edu/research-trials/professional/open-science/ or through a collaboration with a principal investigator affiliated with the UCSF Memory and Aging Center.

## Acknowledgments

Funding support includes grants NIA R01AG060393, the Medical Scientist Training Program T32GM007618, and F30 Fellowship 1F30AG079504-01. Support for the UCSF MAC/ADRC was provided by grants NIA P50-AG023501, P30-AG062422, and P01-AG019724. We thank the Information Commons and Research Analytics Environment teams for access and support with the UCSF EHR data. We acknowledge the use of resources developed and supported by the UCSF IT Academic Research Systems and the UCSF Bakar Computational Health Sciences Institute Information Commons groups, and we thank all members of these groups for their technical support.

