## Supplement for "Characterizing Dementia Phenotypes from Unstructured EHR Notes with Generative AI and Interpretable Machine Learning"

### Extended Methods

#### Filtering of clinical notes

Notes were selected based on specialties related to neurodegeneration care.

This includes notes with any of:

- **Included specialties:** Neurology, Neuropsychology, Neurosurgery, Neuromuscular Medicine, Psychiatry, Psychology, Epilepsy, Neuroradiology, Geriatric Medicine, Physical Therapy
- **Included providers**: Psychologist, Speech and Language Pathologist, Physical Therapist, Occupational Therapist
- **Included departments**: Neurology, Neurosurgery, Geriatric Medicine

#### Local Model Note Verification Prompt

Given the following EHR note, please determine if it contains any meaningful information regarding the following topics:

1. Manual codes (ICD, DSM)

2. Clinical findings

3. Measurements (tests, exams)

4. Medications

5. Medical history

Return your answer as a JSON object with boolean values for the keys:

'manual_codes', 'clinical_findings', 'measurements', 'medications', 'medical_history'.

For example:

{"manual_codes": true, "clinical_findings": false, "measurements": true, "medications": false, "medical_history": true}

#### Local Model Note Verification JSON Schema Validation with Pydantic

{

"properties": {

"manual_codes": {

"title": "Manual Codes",

"type": "boolean"

},

"clinical_findings": {

"title": "Clinical Findings",

"type": "boolean"

},

"measurements": {

"title": "Measurements",

"type": "boolean"

},

"medications": {

"title": "Medications",

"type": "boolean"

},

"medical_history": {

"title": "Medical History",

"type": "boolean"

}

},

"required": [

"manual_codes",

"clinical_findings",

"measurements",

"medications",

"medical_history"

],

"title": "RelevantNote",

"type": "object"

}

#### Full LLM Concept Extraction Prompt

You are a clinical note analyzer specializing in neurological and neuropsychiatric assessments.

TASK:

Analyze clinical notes to extract:

1. Diagnostic codes and references

2. Clinical findings (symptoms and signs)

3. Structured measurements and quantitative data

4. Medications and their details

5. Medical history and past conditions

KEY RULES:

- Follow the provided JSON schema exactly

- Include all diagnostic codes with source systems

- Document both positive and negative findings

- Use EXACT category names as specified below

CATEGORIES:

- Learning

- Memory

- Language

- Executive

- Attention

- Perception

- Motor

- Arousal

- Social

- Valence

- Mood

- Affect

- Behavior

- Thought Content

- Basic Motor

- Complex Motor

- Praxis

- Sensory

- Basic Activities

- Daily Life Activities

- Instrumental Activities

- Sleep

- Endocrinology

- Cardiology

- Pulmonology

- Infectious Disease

- Hematology

- Oncology

- Rheumatology

- Gastroenterology

- Nephrology

- Dermatology

- Urology

- Obstetrics and Gynecology

- Ophthalmology

- Otolaryngology

- Immunology

- Anesthesiology

- Radiology

- Pathology

- Family Medicine

- Internal Medicine

- Physical Therapy

- Occupational Therapy

- Pain Medicine

- Genetics

- Allergy

- Other

DIAGNOSTIC CODE GUIDELINES:

CAPTURE ALL:

- ICD-10 codes

- DSM-5 codes

- Other standardized diagnostic codes

- Provisional and confirmed diagnoses

CRITICAL RULES:

1. Each diagnostic code must be a separate entry

2. Include the coding system (e.g., ICD-10, DSM-5)

3. Document both primary and secondary diagnoses

4. Include supporting quotes as evidence

5. Note diagnostic status (provisional/confirmed/rule-out)

CLINICAL FINDING GUIDELINES:

CAPTURE ALL:

- Signs and symptoms

- Physical examination findings

- Behavioral observations

- Patient-reported symptoms

- Changes from baseline

CRITICAL RULES:

1. Each finding must be a separate entry

2. Include temporal information when available

3. Document severity and frequency if mentioned

4. Note if acute or chronic

5. Include supporting quotes as evidence

MEASUREMENT GUIDELINES:

CAPTURE ALL:

- Complete test scores (e.g., "MMSE 12/30")

- Partial scores and subscores (e.g., "Delayed recall 0/3")

- Timed measures (e.g., "7 animals in 60 seconds")

- Error counts and rule violations

- Discontinued or incomplete tests

CRITICAL RULES:

1. Each finding must be a separate entry

2. Include exact quotes as evidence

3. Use only listed categories

4. Include all contextual details

MEDICATION GUIDELINES:

CAPTURE ALL:

- Current medications

- Recently discontinued medications

- Modified medication regimens

- PRN (as needed) medications

CRITICAL RULES:

1. Each medication must be a separate entry

2. Include complete dosing information when available

4. Note medication status (active/discontinued/modified)

5. Include supporting quotes as evidence

MEDICAL HISTORY GUIDELINES:

CAPTURE ALL:

- Past medical conditions and diagnoses

- Chronic conditions

- Previous surgeries or procedures

- Family history when relevant

- Previous treatments and their outcomes

CRITICAL RULES:

1. Each condition must be a separate entry

2. Include onset/diagnosis timing when available

3. Note current status (active/resolved/chronic/intermittent)

4. Document treatment history

5. Include supporting quotes as evidence

VALIDATION RULES:

1. Categories must match EXACTLY (case-sensitive) from the options below

2. Each finding requires supporting evidence quotes

3. Confidence levels: "high", "medium", "low"

4. Never combine multiple findings

5. Each finding uses ONE category only

Please provide your analysis in valid JSON format.

#### Full LLM Concept Extraction JSON Schema Validation with Pydantic

{

"$defs": {

"Category": {

"enum": [

"Learning",

"Memory",

"Language",

"Executive",

"Attention",

"Perception",

"Motor",

"Arousal",

"Social",

"Valence",

"Mood",

"Affect",

"Behavior",

"Thought Content",

"Basic Motor",

"Complex Motor",

"Praxis",

"Sensory",

"Basic Activities",

"Daily Life Activities",

"Instrumental Activities",

"Sleep",

"Endocrinology",

"Cardiology",

"Pulmonology",

"Infectious Disease",

"Hematology",

"Oncology",

"Rheumatology",

"Gastroenterology",

"Nephrology",

"Dermatology",

"Urology",

"Obstetrics and Gynecology",

"Ophthalmology",

"Otolaryngology",

"Immunology",

"Anesthesiology",

"Radiology",

"Pathology",

"Family Medicine",

"Internal Medicine",

"Physical Therapy",

"Occupational Therapy",

"Pain Medicine",

"Genetics",

"Allergy",

"Other"

],

"title": "Category",

"type": "string"

},

"ClinicalFinding": {

"properties": {

"finding": {

"description": "Specific clinical symptom or sign using standard medical terminology (e.g., 'Anterograde amnesia', 'Gait ataxia')",

"title": "Finding",

"type": "string"

},

"category": {

"$ref": "#/$defs/Category",

"description": "Must be an exact match to one of the predefined neurological/neuropsychiatric categories"

},

"temporal_info": {

"description": "When symptoms started and how they've changed over time (e.g., 'Gradual onset 6 months ago')",

"title": "Temporal Info",

"type": "string"

},

"severity": {

"description": "Description of symptom severity and functional impact (e.g., 'Severe, prevents independent ADLs')",

"title": "Severity",

"type": "string"

},

"evidence": {

"description": "Direct quotes from the text that document this finding",

"items": {

"type": "string"

},

"title": "Evidence",

"type": "array"

},

"confidence": {

"enum": [

"high",

"medium",

"low"

],

"title": "Confidence",

"type": "string"

}

},

"required": [

"finding",

"category",

"temporal_info",

"severity",

"evidence",

"confidence"

],

"title": "ClinicalFinding",

"type": "object"

},

"DiagnosticCode": {

"properties": {

"code": {

"description": "Complete diagnostic code including all characters and modifiers (e.g., 'F03.90', '331.0')",

"title": "Code",

"type": "string"

},

"manual": {

"description": "Source diagnostic manual system - must be one of: ICD-9, ICD-10, DSM-IV, or DSM-5",

"enum": [

"ICD-9",

"ICD-10",

"DSM-IV",

"DSM-5"

],

"title": "Manual",

"type": "string"

},

"diagnosis": {

"description": "Complete diagnostic term as found in the manual (e.g., 'Major Neurocognitive Disorder')",

"title": "Diagnosis",

"type": "string"

},

"status": {

"description": "Whether this is a primary, secondary, or rule-out diagnosis",

"title": "Status",

"type": "string"

},

"evidence": {

"description": "Direct quotes from the text that support this diagnostic code",

"items": {

"type": "string"

},

"title": "Evidence",

"type": "array"

},

"confidence": {

"enum": [

"high",

"medium",

"low"

],

"title": "Confidence",

"type": "string"

}

},

"required": [

"code",

"manual",

"diagnosis",

"status",

"evidence",

"confidence"

],

"title": "DiagnosticCode",

"type": "object"

},

"MedicalHistory": {

"properties": {

"condition": {

"description": "Specific medical condition or diagnosis (e.g., 'Type 2 Diabetes Mellitus')",

"title": "Condition",

"type": "string"

},

"onset": {

"description": "When condition began or was diagnosed (e.g., '2019', '5 years ago')",

"title": "Onset",

"type": "string"

},

"status": {

"description": "Current status of the condition in patient's health",

"enum": [

"active",

"resolved",

"chronic",

"intermittent",

"non-informed"

],

"title": "Status",

"type": "string"

},

"treatment_history": {

"description": "Summary of past and current treatments for this condition",

"title": "Treatment History",

"type": "string"

},

"evidence": {

"description": "Direct quotes documenting this historical condition",

"items": {

"type": "string"

},

"title": "Evidence",

"type": "array"

},

"confidence": {

"enum": [

"high",

"medium",

"low"

],

"title": "Confidence",

"type": "string"

}

},

"required": [

"condition",

"onset",

"status",

"treatment_history",

"evidence",

"confidence"

],

"title": "MedicalHistory",

"type": "object"

},

"Medication": {

"properties": {

"name": {

"description": "Complete medication name including formulation if specified (e.g., 'Donepezil HCl')",

"title": "Name",

"type": "string"

},

"dosage": {

"description": "Complete dosage information including strength and form (e.g., '10 mg tablet')",

"title": "Dosage",

"type": "string"

},

"frequency": {

"description": "How often medication is taken (e.g., 'Once daily', 'Twice daily PRN')",

"title": "Frequency",

"type": "string"

},

"status": {

"description": "Current status of this medication in treatment plan",

"enum": [

"Active",

"Discontinued",

"Modified"

],

"title": "Status",

"type": "string"

},

"evidence": {

"description": "Direct quotes documenting this medication information",

"items": {

"type": "string"

},

"title": "Evidence",

"type": "array"

},

"confidence": {

"enum": [

"high",

"medium",

"low"

],

"title": "Confidence",

"type": "string"

}

},

"required": [

"name",

"dosage",

"frequency",

"status",

"evidence",

"confidence"

],

"title": "Medication",

"type": "object"

},

"StructuredMeasurement": {

"properties": {

"value": {

"description": "Exact numerical score or measurement value (e.g., '23/30', '4 words')",

"title": "Value",

"type": "string"

},

"unit": {

"description": "Unit of measurement or scoring system (e.g., 'Percentile', 'Points', 'Seconds', 'Words per minute')",

"title": "Unit",

"type": "string"

},

"measure_type": {

"enum": [

"Complete test score",

"Partial score",

"Timed measure",

"Discontinued test"

],

"title": "Measure Type",

"type": "string"

},

"test_name": {

"description": "Full standardized name of the test or measure (e.g., 'Mini-Mental State Examination')",

"title": "Test Name",

"type": "string"

},

"test_domain": {

"description": "Cognitive/behavioral domain being tested (e.g., 'Memory', 'Executive function')",

"title": "Test Domain",

"type": "string"

},

"evidence": {

"description": "Direct quotes containing the measurement data",

"items": {

"type": "string"

},

"title": "Evidence",

"type": "array"

},

"confidence": {

"enum": [

"high",

"medium",

"low"

],

"title": "Confidence",

"type": "string"

}

},

"required": [

"value",

"unit",

"measure_type",

"test_name",

"test_domain",

"evidence",

"confidence"

],

"title": "StructuredMeasurement",

"type": "object"

}

},

"properties": {

"diagnostic_codes": {

"description": "All diagnostic codes mentioned in the note",

"items": {

"$ref": "#/$defs/DiagnosticCode"

},

"title": "Diagnostic Codes",

"type": "array"

},

"clinical_findings": {

"description": "All clinical symptoms and signs documented",

"items": {

"$ref": "#/$defs/ClinicalFinding"

},

"title": "Clinical Findings",

"type": "array"

},

"structured_measurements": {

"description": "All test scores and quantitative measurements",

"items": {

"$ref": "#/$defs/StructuredMeasurement"

},

"title": "Structured Measurements",

"type": "array"

},

"medications": {

"description": "All medications mentioned with their details",

"items": {

"$ref": "#/$defs/Medication"

},

"title": "Medications",

"type": "array"

},

"medical_history": {

"description": "All historical and current medical conditions",

"items": {

"$ref": "#/$defs/MedicalHistory"

},

"title": "Medical History",

"type": "array"

}

},

"required": [

"diagnostic_codes",

"clinical_findings",

"structured_measurements",

"medications",

"medical_history"

],

"title": "AnalysisResult",

"type": "object"

}

### Best parameters identified from HDBScan on embedded concepts

Best Parameters:

'min_cluster_size': 50

'min_samples': 35

'cluster_selection_epsilon': 0.0

with silhoutte score of 0.5491373.

51 unique clusters identified, excluding noise.

### Supplemental Table 1: Example of 6 symptom clusters with example text

| **Cluster** | **LLM-assigned Label** | **# of findings** | **Example Text** |
| --- | --- | --- | --- |
| 1 | Masked Facies and Reduced Blink Rate | 52 | - masked facies  - presentation was notable for very masked facies and a stiff wide-eyed stare throughout the examination  - [pt] was described as having marked psychomotor slowing with masked facies and bradyphrenia  - other findings on examination include masked facies with reduced blink frequency  - [pt] facial expression is less animated than previous |
| 2 | Acalculia and Calculation Impairment | 71 | - [pt] has trouble with calculations which is highly unusual for him  - difficulty solving mathematical calculations  - [pt] evidenced difficulty on a screen of calculations missing points on a complex multiplication problem and refusing to attempt two double-digit subtraction and addition problems  - [pt] also had difficult with calculations  - calculations were also impaired both for multiplication and subtraction |
| 3 | Cardiac Rhythm and Blood Pressure Variations | 84 | - patient carries a diagnosis of orthostatic hypotension  - [pt] was not clinically orthostatic  - on general physical examination [pt] was tachycardic  - mildly elevated potassium 56  - pulse 41 |
| 4 | Spinal Degenerative Changes | 110 | - normal bone mineralization vertebral body heights and intervertebral disc space are well-maintained at l3-l5  - there is sclerosis of the superior aspect of both sacroiliac joints  - corresponding to the areas of signal abnormality and enhancement on the mri performed [on date] c7 t1 and t4 vertebral bodies there is evidence for subtle radiolucency with vertical trabecular thickening  - l4-l5 vacuum phenomenon in the disk and disk space narrowing no significant canal or neural foraminal narrowing moderate bilateral facet arthropathy  - degenerative changes of the thoracic spine are noted |
| 5 | Urinary and Bowel Incontinence | 251 | - [pt] is incontinent of urine frequently  - [pt] is continent but has had intermittent constipation with loose bowels occurring every 3 days  - [pts] daughters say that [pts] bowel movements are irregular and that [pt] is now requiring a new laxative  - [pt] lost bowel/bladder control  - [pt] is incontinent so wears a depends at all times |
| 6 | Poor Judgment and Social Conduct Issues | 57 | - [pts] insight is poor [pts] judgment is poor [pt] is not reliable in the history frequently changing [pts] story  - [pt] exhibited uncharacteristically poor judgment in not bringing her to the hospital until prompted by other family members  - poor judgment  - poor judgment and decision making--accumulating credit card debt constantly worry about everything repeating [pt] unresolved concern that [pt] and [pt’s] family members will be going to jail due to credit card debt  - poor judgment |
